# Operationalizing a complex acute clinical trial: Lessons from the BEACH study

**DOI:** 10.1101/2025.03.28.25324776

**Authors:** Gracey Sorensen, Will Remillard, Maia Schlechter, Michael Kampp, Lauren H. Sansing, Cailin Whisler Brady, Kaley Kidahl, Wendy Ziai, Linda Van Eldik, Ashley Distasio, Jing Lu, Jessica Magid-Bernstein, Dan Hanley

**Affiliations:** Department of Neurology, Yale School of Medicine, New Haven, CT; Department of Immunobiology, Yale School of Medicine, New Haven, CT; Division of Brain Injury Outcomes, Johns Hopkins School of Medicine, Baltimore, MD; Department of Anatomy and Neurobiology, University of Kentucky, Lexington, KY; Yale New Haven Hospital, New Haven CT; Investigational Drug Service, Yale New Haven Hospital, New Haven, CT

## Abstract

**Purpose:** To outline the workflow, challenges, and key roles involved in operationalizing a complex, acute clinical trial protocol requiring multidisciplinary collaboration.

**Summary:** Yale University School of Medicine and the Neuroscience Intensive Care Unit (NICU) at Yale New Haven Hospital leverage interdisciplinary collaboration to successfully enroll patients into complex clinical trials, including the Biomarker and Edema Attenuation in IntraCerebral Hemorrhage (BEACH) trial (ClinicalTrials.gov identifier: NCT05020535). The research team proactively educates nursing staff and clinicians on the study protocol, in addition to facilitating communication with the investigational drug pharmacy and supporting treating physicians in screening, enrollment, and follow-up. With 24/7 access to the research team, study coordinators are present for all protocol steps, including test article infusion, biospecimen collection and processing, and participant interactions. Successful execution of the BEACH trial relies on five key domains: ensuring patient safety in a high acuity setting, optimizing screening and enrollment processes, implementing efficient workflows for pharmacokinetic sampling and test article administration, identifying signals of efficacy, and adapting to operational challenges. These domains require precise coordination, clear communication, and adaptability within dynamic patient care environments. By streamlining workflows and maintaining open communication, the research team enhances efficiency and optimizes patient enrollment while upholding the highest standards of ethical research and patient care.

**Conclusion:** Implementation of the BEACH trial at Yale exemplifies the critical role of interdisciplinary collaboration in advancing clinical research. By integrating research into patient care, the study team not only enhances trial efficiency, but also contributes to the development of innovative treatment strategies for intracerebral hemorrhage. Moving forward, the lessons learned from operationalizing BEACH can inform best practices for future acute trials, ensuring that research continues to drive meaningful improvements in patient outcomes.

## Introduction

Successful clinical trial execution relies on a well-structured workflow and seamless collaboration among multidisciplinary teams. At Yale School of Medicine, the divisions of Vascular Neurology and Neurocritical Care maintain a 24/7 core research team composed of six research postgraduates, a manager, and faculty investigators. This team is actively involved in all aspects of trial operations, from initial screening and enrollment to follow-up and data management.

Collaboration between study coordinators, Principal Investigators (PIs), clinical teams, nursing staff, and the investigational pharmacy is essential to ensure efficient and accurate trial execution. Effective coordination allows the team to navigate both expected and unforeseen challenges, ensuring protocol adherence while maintaining high-quality patient care.

This article aims to provide hospital research centers with a comprehensive understanding of the workflow, challenges, and key roles involved in executing a clinical trial that requires significant nursing participation and multidisciplinary collaboration.^1^ Using the Biomarker and Edema Attenuation in Intracerebral Hemorrhage (BEACH) trial at Yale New Haven Hospital’s Neuroscience Intensive Care Unit (NICU) as a case study, we outline the methods used to adhere to a rigorous study protocol. The discussion is organized into five critical domains of clinical trial execution:

1. **Safety** – Ensuring patient well-being and protocol adherence in a high-acuity setting.
2. **Screening and Enrollment** – Identifying eligible patients and overcoming logistical barriers.
3. **Acquiring Pharmacokinetics** – Strategies for precise and timely sample collection and test article administration.
4. **Signals of Efficacy** – Measuring and interpreting early indicators of therapeutic impact.
5. **Adapting to Unexpected Changes** – Managing operational challenges and protocol deviations.

### BEACH Clinical Trial Overview

BEACH is a Phase II, multicenter, double-blinded, randomized, placebo-controlled trial evaluating the use of an intravenous (IV)-infusion of MW-189 (referred to as test article), or placebo in patients diagnosed with spontaneous intracerebral hemorrhage (ICH).^2^ Inclusion criteria include but are not limited to diagnosis of a spontaneous, non-traumatic ICH, symptom onset within 24-hours of presentation, 5-60cc intraparenchymal bleed volume, and no neurosurgical interventions outside of placement of an external ventricular drain. Randomization occurs on a central website, assigning patients 1:1 to test article or placebo. Per study protocol, the first infusion must be within 24 hours of onset of the patient’s symptoms. The study involves 10 infusions (of test article or placebo), once every 12 hours, for a duration of 5 days. There are 6 electrocardiograms (ECGs), 16 blood sample collections, 20 vital signs collections, and brain imaging timed around the infusions (Figure 1). Follow-ups include constant monitoring for adverse events, a day-30 in-person visit, a day-90 remote visit, and a day-180 in-person visit. Given the complexity of the trial protocol, BEACH requires constant collaboration between research coordinators, the clinical team, nursing staff, and pharmacists for a successful enrollment.

**Figure 1.**
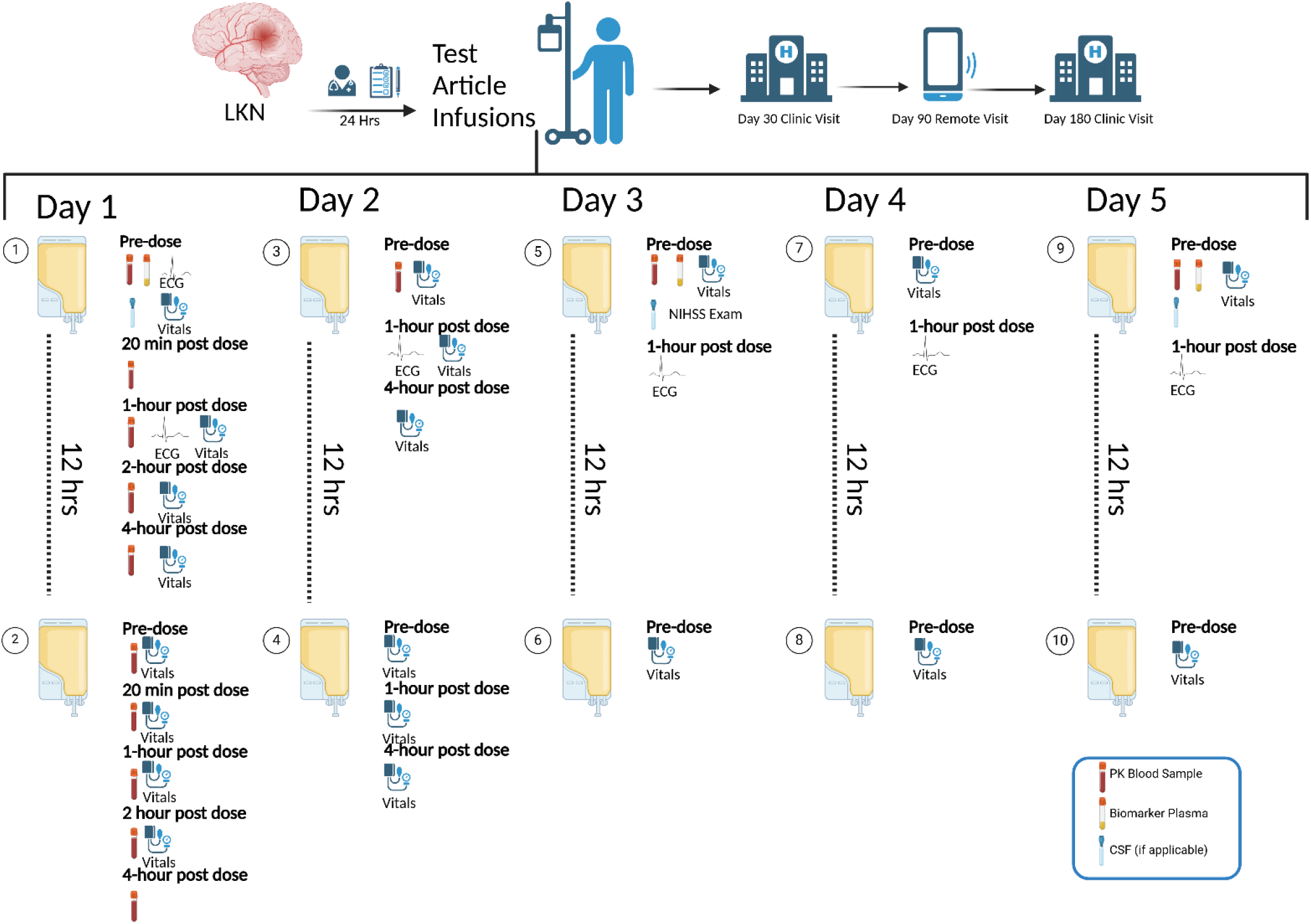
BEACH Protocol Flow Diagram. Schematic representation of the BEACH trial protocol, showing key steps from screening and randomization to in-hospital infusions and long-term follow-up. Figure created with BioRender

### Yale Post-Graduate Clinical Research Team

The Yale Department of Neurology Divisions of Vascular Neurology and Neurocritical Care employ a team of post-graduate Clinical Research Associates, who serve as study coordinators for the BEACH trial and other acute brain injury trials. Using a rotating on-call schedule, the study coordinators share a research phone, providing the clinical team with 24/7 access to the on-call coordinator for screening and enrollment assistance. The position operates on a two-year cycle, with three new postgraduates hired annually. This structured rotation ensures continuity in trial operations, as new members are consistently trained to enroll patients, minimizing disruptions and maintaining seamless study execution.

#### 1. Safety -data collection/primary outcome

As a Phase II trial, the primary outcome of the BEACH clinical trial is all-cause mortality within the first seven days post-randomization. Secondary outcomes include 30-day all-cause mortality, hematoma expansion and recurrent ICH, brain infection, pharmacokinetic parameters, radiographic measures of edema, and inflammatory and neuronal injury biomarkers in plasma and cerebrospinal fluid during and at the end of treatment (days 1–5).^2^ Ensuring patient safety is the most critical factor in subject enrollment, and this requires extensive nursing education by study coordinators both prior to trial initiation and in real time when a subject is enrolled.

One of the primary challenges in maintaining compliance is the continual onboarding of new faculty members, fellows, and nurses, who must be trained in trial procedures. To mitigate protocol deviations, structured training programs and regular refresher sessions have been implemented, significantly improving adherence over time (Figure 2). Additionally, a nightly reminder text is sent to the clinical team by study coordinators, ensuring that acute screening remains a priority.

**Figure 2.**
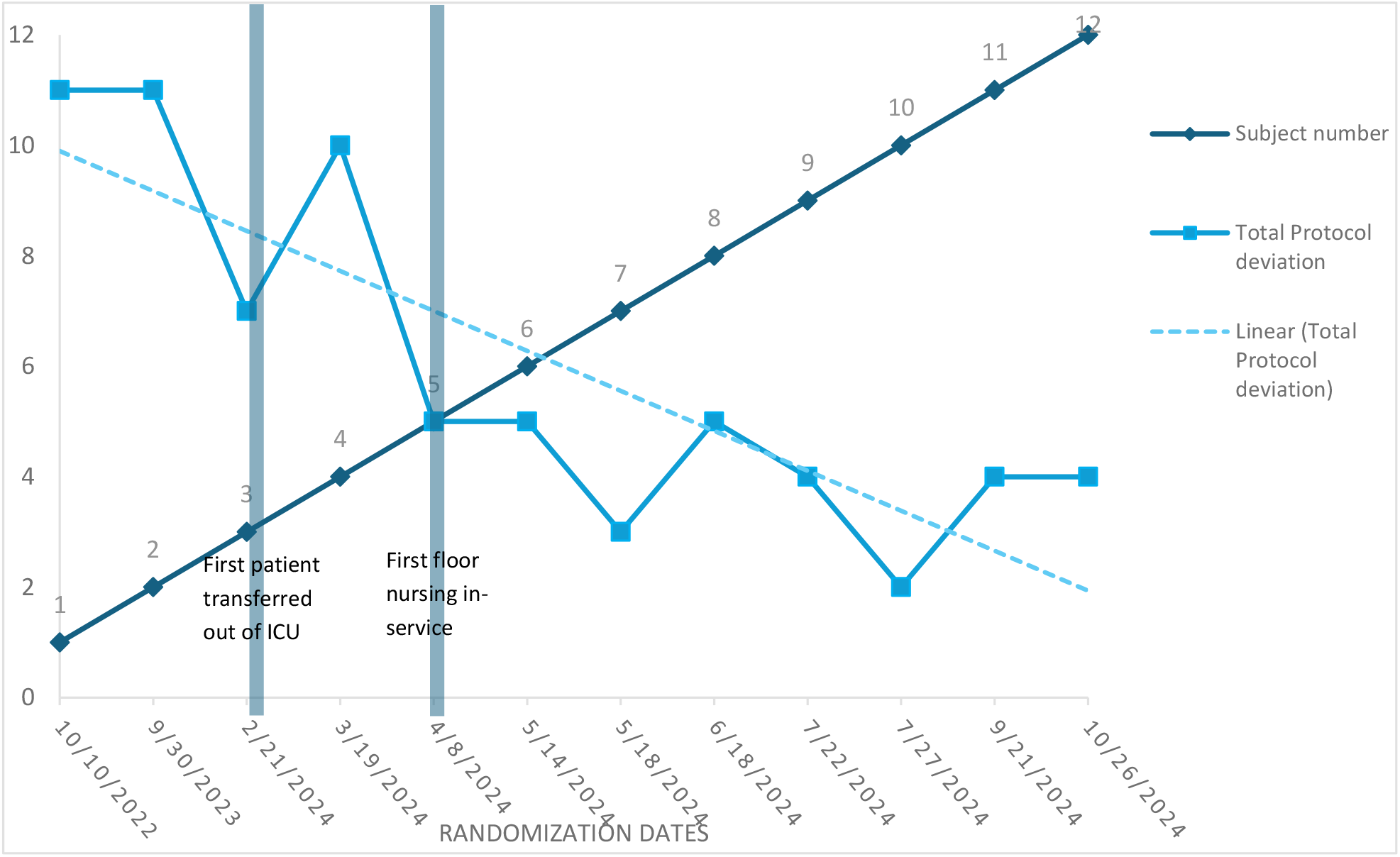
Yale BEACH subject randomization dates and protocol deviations. This figure shows a decrease in protocol deviations as the number of BEACH trial subjects increases, highlighting a strong negative correlation with fewer deviations as enrollment reaches 12 subjects. Notably, subject 3 was the first to transfer out of the ICU during infusions (highlighted in blue), and the first nursing in-service occurred on the floor on April 3, 2024 (highlighted in blue). Subjects 3-9, 11, and 12 all had infusions completed in both the ICU and the stepdown unit.

### Education and training

#### Central Site Training

Sites must complete initial training designated by the central site, Johns Hopkins University, prior to initiating enrollment. All involved staff must complete training on Biospecimen Collection, Study Drug Administration, Protocol & Safety, and Neuroimaging. Each site designates who will be trained, and at Yale, all NICU and Stroke Service attendings and fellows complete the full training. Once completed, the central site grants approval to enroll. Updated training is available on a central website in the event of protocol modifications.

#### Site specific nurse education

Given the critical role of nurses in trial enrollment and execution, the Yale clinical research team provides both proactive and on-the-spot training. Study coordinators meet monthly with the NICU nurse educator and nurse manager to refine trial workflow. Additionally, ad hoc in-service sessions are held for ICU and stepdown unit nursing staff to review protocol updates and build collaboration between nursing staff and study coordinators. During enrollment, coordinators engage nursing staff early to provide real-time training on infusion, safety, and protocol timing.

#### Resources

- **Nursing-One-Sheet –** Displays calendar of events, nursing tasks, contact information, and infusion safety information.
- **Patient Room Calendar** – Displays infusion schedules, vitals, blood draws, CSF collection, ECGs, and imaging timelines.
- **QR Code** – Links to the research website with trial details, inclusion/exclusion criteria, and study methods.
- **Electronic Medical Record (EMR)** – Stores signed consent forms and includes a **treatment team sticky note**, providing an updated study overview, 24/7 research team contacts, PI details, and infusion schedule.

Early subjects enrolled saw increased protocol deviations when patients were transferred to floors with untrained staff (Figure 2). To address this, designated units with trained nurses are prioritized for BEACH participants. When transfers to these units are not possible, study coordinators proactively engage the appropriate charge nurse and nurse managers to facilitate training and preparation.

### Central site communication

BEACH trial sites maintain frequent communication with the central site, Johns Hopkins University. Biweekly calls address site-specific challenges and procedural adherence, while monthly webinars review enrollment, reinforce protocol compliance, and provide updates from the global PIs. An online database is utilized to upload all patient information and to alert sites via email to promptly correct data entry errors.

### Adverse event reporting

A study coordinator attends daily rounds on BEACH subjects throughout enrollment, ensuring prompt reporting of adverse events, protocol deviations, administration of prohibited medications, and unexpected events to the central site. Daily collaboration with the clinical team ensures clarity on adverse events and treatment plans.

#### 2. Screening and Randomization

Given the trial’s acute nature and the requirement for initiation of infusion within 24 hours of symptom onset, early identification and rapid enrollment of eligible patients are crucial. Efficient screening, coordination with the clinical team, and clear communication ensure timely intervention. Yale’s dedicated team of study coordinators is available 24/7 to facilitate screening and enrollment. The process begins with automated alerts from Viz.ai and Yale-generated head bleed notifications for patients presenting to the emergency department with a brain hemorrhage.

### Head Bleed Alerts and the Research Team Email

Internal Yale head bleed alert notifications are sent to the 24/7 research phone and the study PI, prompting immediate screening initiation. Additionally, each morning, study coordinators review our internal research team email, which compiles details on newly admitted stroke and NICU patients, including diagnoses, treatments, and imaging results. These steps ensure no eligible patients are overlooked.

### Ensuring accurate bleed measurement

Accurate bleed measurement is essential for determining eligibility in the BEACH trial, which requires an intraparenchymal hematoma volume of 5-60cc. The BEACH trial integrates the Viz RECRUIT platform, an AI-driven tool that automates computed tomography (CT) scan analysis to assist in identifying potential participants.^3^ At Yale, Viz.ai provides real-time mobile notifications of all CT scans with ICH volume measurements that meet BEACH inclusion criteria, including those completed at Yale-affiliated hospitals. These automated alerts notify the study PI and study coordinators about new patients who may meet enrollment criteria, expediting the screening process. For accuracy, volumes can also be calculated manually using the ABC/2 method^4^ or assessed by a central site. Any discrepancies are resolved with the local or global PI to ensure the most representative measurement.

### Eligibility Confirmation

Upon receiving an alert, the study coordinator assesses eligibility and consults the clinical team and study PI for confirmation and clarification on any details which may be unclear on initial chart review. Throughout the process, the on-call coordinator maintains continuous communication with the clinical team and study PI via the 24/7 research phone, and the global PIs are available via the BEACH hotline for any questions regarding eligibility.

### Consent

Once a patient is determined to be eligible for enrollment, the study coordinator arranges with the clinical team to have a consent conversation with the patient and/or their legally authorized representative. The stroke or NICU faculty member or fellow introduces the trial, answers any clinical or research questions that arise, and obtains consent. The study coordinator is present throughout the conversation to assist with any additional study-specific or logistical questions that may come up.

### Randomization

After confirming eligibility and obtaining consent, the coordinator enters screening data into the central website and uploads the signed consent form. The system generates randomization details, which are sent directly to the Yale’s Investigational Drug Service (IDS) to preserve the trial’s double-blind integrity.

### Enrollment Preparation

Once randomized, the research team coordinates with the IDS and nursing staff to schedule the first infusion and estimates the timing for the subsequent nine doses. The clinical team ensures adequate blood access (via arterial line or midline catheter), and the research coordinators provide on-the-spot reminders regarding prohibited medications and nursing requirements.

#### 3. Acquiring pharmacokinetics

Precise pharmacokinetic data collection is critical to understanding the safety, tolerability, and potential efficacy of MW189 in the BEACH trial. At Yale, a comprehensive, multidisciplinary strategy has been established to ensure that every step from drug preparation to lab processing is executed with accuracy.

### Pharmacy Coordination

Given the acute nature of the BEACH trial, the 24/7 pharmacy at Yale New Haven Hospital (YNHH) plays a critical role in ensuring the timely preparation and delivery of the test article in a double-blinded manner. To ensure efficiency, IDS is notified immediately upon identification of a potential participant, even before consent is obtained. Notification occurs via the pharmacy’s on-call research phone and email, enabling timely preparation of the test article. This proactive approach ensures that the test article is readily available for administration without delay. To uphold the double-blind nature of the trial, the test article is packaged in a brown paper bag with an amber-covered sheathing over the IV bag, as the test article and placebo differ in color. Once the test article is prepared, IDS notifies the on-call study coordinator, who then retrieves and delivers it to the patient’s room. A scannable QR code on the exterior bag provides drug information and study HIC number without revealing the test article contents. This process is documented in the medication drug data sheet to track the test article, prevent misplacement, and ensure timely administration. The study coordinator coordinates with IDS to pick up the test article for all administrations and deliver them directly to the patient’s nurse.

### Lab Draws and Processing

Given the trial’s intensive sampling schedule, site protocol requires NICU admission with established vascular access for blood draws (arterial line or midline catheter) for at least the first 24 hours after randomization (two infusions). This ensures rapid, minimally invasive blood draws for accurate pharmacokinetic assessments by trained nurses. Study coordinators, trained in lab processing, aliquot and store all samples within a strict time frame. During transport to the YNHH Hospital Research Unit for processing, samples are kept on ice in a designated biological sample container.

### Collaboration with Nursing on Timing

Effective communication with nursing staff is essential to synchronize blood draws with drug infusions (Figure 1). Infusion times are carefully scheduled to avoid nursing shift changes, which occur at 07:00 and 19:00. Nurses prepare for each infusion by setting up the IV line to ensure that the system is in place before the pre-infusion blood draw. This coordination guarantees that sample collection occurs within the designated time windows, thereby minimizing protocol deviations and ensuring high-quality pharmacokinetic data. Study coordinators are present for all blood draws to assist with timing, document collection times, and confirm adherence to the strict five-minute collection window.

### Infusion Planning and Set-Up

Before each infusion, detailed planning is guided by a site-specific manual of procedures developed by the study coordinators. Infusion set-up involves ensuring all necessary equipment, including IV tubing, amber blinding sheathing, tape, scissors, labeled collection tubes, and a transport container with ice, is properly arranged in the patient’s room. Timing coordination is critical, as the initial infusion establishes the schedule for all subsequent sdoses and sample collections. Pre-infusion checks require close collaboration with pharmacy and nursing staff to confirm that pre-infusion lab samples are drawn immediately after setup is complete. Once collected, study coordinators record the time on the patient’s calendar, which remains in the patient’s room, and update the electronic medical record to ensure proper documentation and adherence to protocol. From the first infusion, all subsequent infusions are timed and communicated between nurses at change of shift, to ensure protocol adherence.

#### 4. Signals of efficacy

To obtain signals of efficacy within a clinical trial, patient outcomes and samples must be collected within the correct window. In the BEACH clinical trial, coordinators begin scheduling follow-ups early to determine the patient’s discharge location and whether the visit will be conducted in person or remotely.

### Follow-ups

Patient follow-ups are scheduled based on the availability of both the patient and the PI. Follow-ups occur in-person at day 30 post-ICH, remotely via phone at day 90 post-ICH, and in-person again at day 180 post-ICH. In-person follow-up appointments are typically held at the Hospital Research Unit at YNHH.

For patients discharged to a rehabilitation facility, coordinators contact the facility at the time of discharge to ensure awareness of the required follow-ups for trial participation. If the patient remains at the rehabilitation facility on the scheduled follow-up dates, coordinators arrange a remote visit with assistance from facility staff or family members. To minimize disruption to the patient’s rehabilitation, coordinators collaborate with the care team to schedule visits around therapy sessions, such as occupational or physical therapy.

When possible, in-person follow-up appointments may also be conducted at outside rehabilitation facilities. For patients with transportation difficulties, coordinators arrange transportation to the YNHH Hospital Research Unit to ensure the follow-up visit is completed.

### Biospecimen Collection and Shipping

Study coordinators are responsible for storing and shipping all biospecimens to the central lab for analysis. Samples are stored in a -80°C research freezer, and there is access to 24/7 dry ice for shipping. Before shipment, coordinators confirm with the central lab that laboratory staff will be available to process the samples, which are then shipped overnight on dry ice.

#### 5. Adapting to unexpected changes and unanticipated trial operations

Due to the extended duration of infusions and the varying stability of enrolled patients, many patients are managed by multiple clinical teams and require a complex workup during their hospitalization. This presents challenges in ensuring that all clinical team members are informed of essential infusion details and aware of prohibited medications.

### Transferring between floors

During the five-day infusion, patients frequently transition from the NICU to the stroke stepdown unit, posing challenges in maintaining adherence to the BEACH protocol with new nursing staff. This challenge arises because NICU nurses, who typically manage only two patients, are more familiar with acute clinical trial operations and protocols, whereas stepdown unit nurses oversee a larger patient load and may have less familiarity with the inpatient clinical trials.

To facilitate a smooth transition, the charge nurse on the stepdown unit is notified in advance of the plan for an enrolled patient to be transferred out of the ICU. Study coordinators at YNHH meet with the nurse manager to identify staff responsible for lab draws, ECGs, and vitals. Unlike in the ICU, where nurses handle these tasks directly, the stepdown unit relies on additional team members for collection vital signs and ECGs, requiring adjustments in communication and workflow for study coordinators.

The on-call coordinator remains present for each infusion and is available to address any protocol-related questions. As there is a 2-hour window for drug infusion, coordinators arrive early in this window to allow time for additional education for stepdown nurses, if needed. To ensure continuity, all BEACH trial materials accompany the patient when they change rooms, providing stepdown nurses with essential resources, including blood draw and ECG instructions and contact information for the on-call coordinator.

### Drug-Drug Interaction (DDI) Management

Due to the pharmacological properties of MW-189, medications classified as CYP2C9 substrates and CYP3A inhibitors cannot be administered during the five-day infusion period and for 24 hours following the final dose. To ensure patient safety and protocol adherence, addressing potential DDIs requires a coordinated, multi-disciplinary approach involving clinical team training, nurse education, and pharmacy preparation.

The screening process begins with study coordinators identifying any prohibited medications in a patient’s home regimen. They collaborate with the clinical team to determine whether alternative medications can be safely substituted prior to enrollment. For example, if a patient is taking an exclusionary anti-hypertensive medication, the team assesses whether an alternative blood pressure medication can be used during the first 6 days of the patient’s hospitalization to facilitate safe participation in the study.

In addition, coordinators work closely with the pharmacy to integrate DDI alerts into the EMR system. The EMR medication report for the test article includes details on potential DDIs and the contact information for the research team. Nursing staff are instructed to reference both the EMR and in-room patient documentation to ensure awareness of prohibited medications.

During rounds, coordinators reinforce these precautions by reminding the clinical team of restricted medications and reviewing daily orders to prevent the initiation of any excluded drugs. This proactive approach ensures continuous monitoring and mitigates the risk of unintended drug interactions, prioritizing patient safety throughout the study.

## Discussion

The successful execution of the BEACH clinical trial requires meticulous preparation, cross-disciplinary collaboration, and ongoing education. Effective communication between the research team, clinical team, pharmacy, and central coordinating center is essential for adhering to the protocol and ensuring patient safety.

The five domains outlined in this article are essential for maintaining research integrity in an acute clinical trial such as BEACH:

**Domain 1:** Prioritizing rigorous safety protocols, enhancing real-time adverse event monitoring, and advancing nurse education will strengthen adherence, improve patient outcomes, and ensure data integrity.

**Domain 2:** Streamlining AI-assisted screening, accelerating eligibility confirmation, and maintaining 24/7 research team coordination will boost enrollment, minimize missed eligible patients, and ensure timely infusions, reinforcing protocol adherence and trial reliability.

**Domain 3:** Precise test article preparation, coordinated blood draws, and strict timing adherence will enhance data accuracy, ensure compliance, and optimize MW189 pharmacokinetic assessments in the BEACH trial.

**Domain 4:** Proactively scheduling follow-ups, coordinating with rehabilitation facilities, and facilitating transportation will improve patient retention and strengthen the reliability of efficacy assessments.

**Domain 5:** Enhancing communication during transfers, designating trained nursing units, and managing drug-drug interactions will reduce protocol deviations and improve adherence.

At Yale, we have developed clear protocols to address these domains, to ensure patient safety, seamless patient screening and enrollment, strict adherence to drug infusion and biospecimen collection, timely follow up, and effective communication between clinical and research teams. We have learned from each patient enrollment and have adapted our protocols overtime to optimize patient safety and high-quality data collection.

## Conclusion

Through ongoing education, structured workflows, and a commitment to continuous learning, protocol deviations have steadily declined, improving overall trial execution. Each patient enrolled not only contributes to scientific discovery but also strengthens the trial’s operational framework, reinforcing best practices for future research endeavors.

## Data Availability

All data produced in the present work are contained in the manuscript

## Acknowledgement

The BEACH trial’s success is due to the dedication of many individuals and teams. We thank the patients and families for their trust and participation.

We appreciate the support of Yale New Haven Hospital’s NICU, Stroke Team, Emergency Department, and Neurocritical Care faculty. Special thanks to the nursing staff, IDS pharmacy, and research coordinators for ensuring protocol adherence and patient safety.

We also acknowledge Viz.ai and Vision EDC for technological support. Additionally, we thank the Yale Center for Clinical Investigation (YCCI) and the trial’s central coordinating center, Johns Hopkins University, for their guidance and regulatory oversight. Lastly, we thank the principal and sub-investigators for their leadership and mentorship.

## Contributor information

BEACH is directed by the University of Kentucky and the Johns Hopkins University BIOS Clinical Trials Coordinating Center.

The BEACH study and the authors are supported by a grant from the National Institutes of Health National Institute on Aging (R01AG069930, DH & LVE). Additional support came from the National Institutes of Health National Center for Advancing Translational Sciences (U24TR001609, DH).

## Ethical Approval/Informed Consent

The Johns Hopkins Medicine IRB, serving as the single IRB, approved this study. The Yale University IRB approved this study. The trial will be conducted in accordance with the principles of the Declaration of Helsinki, the Good Clinical Practice guidelines of the International Council of Harmonisation, and all applicable regulatory requirements. Written informed consent has been obtained from all participants or their legally authorized representatives.

## Clinical Trial Registration

ClinicalTrials.gov identifier NCT05020535.

